# Sensitive and Specific Early-Stage Breast Cancer Detection using Deep Proteome Profiling from Plasma

**DOI:** 10.64898/2025.12.07.25341735

**Authors:** Alec Horrmann, Yash Travadi, Jacob Carey, Ella Boytim, Kevin Mallery, Grant Schaap, Carissa Rungkittikhun, Kaylee Judith Kamalanathan, Nathaniel R. Bristow, Catalina Galeano-Garces, Adam Groth, Harrison Ball, Alexa R. Hesch, Pooja Advani, Justin Hwang, Badrinath R. Konety, Justin M. Drake

**Affiliations:** Astrin Biosciences, Saint Paul, MN; Department of Medicine, Masonic Cancer Center, University of Minnesota, Minneapolis, MN; Division of Hematology and Oncology, Mayo Clinic, Jacksonville, FL; Allina Health, Minneapolis, MN

**Keywords:** liquid biopsy, proteomics, mass spectrometry, dense breast tissue, early detection, breast cancer

## Abstract

Proteome-guided liquid biopsy tests hold immense promise for the future of early cancer detection. Our previous published work has shown strong performance identifying early-stage breast cancer patients using prospectively collected, case-controlled samples. Here, we analyzed the plasma proteome of 1,259 biobanked samples consisting of healthy women and women with breast cancer. The Astrin Biosciences’ breast cancer early detection test is a laboratory developed test (LDT) that uses a protein-based machine learning classifier to identify breast cancer with high accuracy. The classifier was trained on 845 women, comprising of 466 healthy and 379 with newly diagnosed, treatment naïve breast cancer and validated on 397 women (195 healthy and 202 breast cancer). All plasma samples were processed in a blinded manner coupled with semi-quantitative, label-free mass spectrometry (MS)-based analysis. The validation performance achieved 92.3% specificity, 92.6% sensitivity and an AUC of 0.975. Sensitivity remained high across all breast cancer stages and pathological and molecular subtypes. Gene set enrichment analyses (GSEA) identified epithelial-to-mesenchymal transition (EMT) and PI3K-AKT signaling as enriched in the breast cancer samples, highlighting that our test can identify cancer-related proteins in early-stage patients. A simulated population demonstrates the utility of our test as a supplement to mammography, detecting nearly all (93%) breast cancers missed by mammography and reducing the number of false positives relative to MRI and Contrast-Enhanced Mammography (CEM) by >10-fold. Overall, our proteomic data demonstrates high sensitivity and specificity in women with breast cancer, especially at early stages, and is a favorable supplemental test post mammogram.

## Introduction

Mammography is the gold standard for population-based breast cancer screening. However, its detection sensitivity is significantly reduced in women with dense breast tissue, who face an elevated risk of developing breast cancer^1–4^. Recent updates to the FDA’s mammography regulations^5^ underscore the increasing focus on optimizing screening strategies in this patient population (representing approximately 50% of the US screening population). MRI and Contrast-Enhanced Mammography (CEM) have recently shown potential as a promising supplement to traditional mammography screens in the BRAIDs trial^6,7^. A consensus on the best imaging strategy for individuals with dense breasts has not been established. Although supplemental imaging such as breast MRI can improve cancer detection in women with dense breasts, its use is not routinely recommended for average-risk individuals due to increased rates of false positives, over-diagnosis, additional biopsies, concern for contrast related toxicity, patient anxiety, and cost. Therefore, supplemental MRI imaging is often reserved for use in high-risk patients (with a lifetime risk of breast cancer ≥20%). There is currently a lack of sufficient evidence to justify the benefit of use in patients with high breast density who otherwise have a negative screening mammogram^8^, creating an opportunity for new supplemental screening tests to help bridge this gap for high-risk patients.

The implementation of liquid biopsy-based tests for early cancer detection is poised to be an important component of the supplemental screening landscape, including breast cancer. These tests can help reduce unnecessary imaging or other diagnostic interventions, thus decreasing costs related to current supplemental screening guidelines while increasing detection rates. The low sensitivity for breast cancer at early stages with nucleotide-based tests^9^ offers opportunities to bring new technologies and approaches, such as proteomics, to at-risk women where supplemental screening would typically be implemented. Proteomics is an emerging field in the clinical diagnostics space with niche uses for blood–based diagnostic testing^10–12^. This has typically been through analyses of a single protein or a few proteins. The incorporation of large-scale, unbiased (or shotgun) proteomics using mass spectrometry (MS)-based technologies for clinical diagnostics has not been utilized to date.

Here, we assessed a large cohort of biobanked samples consisting of 1,259 women to detect breast cancer as early as stage 0 through deep proteomic profiling via MS. We have further refined and validated our automated processing pipeline to continuously prepare and analyze hundreds of plasma samples with low sample error rates and high reproducibility while obtaining sensitivities near 90% across all stages of breast cancer using our proprietary spectral library with an improved machine learning algorithm. While this workflow is currently limited to breast cancer detection, this pipeline can be broadly adapted to other types of cancer and diseases.

## Materials and Methods

### Sex as a Biological Variable

Our study examined female subjects only since >99% of breast cancer cases are in females.

### Sample Acquisition and Processing

Breast cancer and healthy plasma samples were purchased from two different biobanks (ProteoGenex and BioIVT) and shipped frozen to Astrin Biosciences. Cancer samples were selected based on the following criteria: 1) treatment naïve, 2) no prior history of cancer, and 3) a confirmed diagnosis of breast cancer based on histopathological analysis of subsequent biopsy or resected tissue samples. Healthy female donors were selected to create an age-matched population.

Samples were prepared as described by Horrmann et al^13^. Briefly, plasma aliquots were loaded onto a Microlab Prep (Hamilton) robot liquid handler along with process and digestion controls and positive and negative controls comprising pooled breast cancer and healthy patient plasma, respectively. A proprietary functionalized superparamagnetic bead solution was added to allow for preferential protein capture while leaving behind contaminating high abundance proteins. The sample plate along with wash and lysis plates were transferred to a Kingfisher Flex (Thermo Fisher) which performed a series of washes ending with sample lysis on a heated shaker. Using the Microlab Prep, the samples were transferred to a 96-well LoBinding plate (Eppendorf, 0030129512) for all subsequent sample steps. A protein recapture buffer was then added to promote protein recovery followed by a series of buffer exchanges ending with digestion buffer (pH 8.0) and digested with Lys-C and trypsin overnight. The next morning, the peptide-containing supernatant was removed and dried on a vacuum concentrator (Labconco).

For plate and mass spectrometry controls, we purchased enolase (Waters), HeLa lysates (Thermo Fisher), and nine different positive and negative controls from a biobanked source. The positive and negative controls were pooled to create a common sample source to assess performance on a plate-by-plate basis. Multiple blank controls were also included with each plate to test both for sample carryover and any contamination throughout the sample processing.

### Characterization of breast cancer molecular subtypes

Typical subtyping requires pathology reports that include ER/PR/HER2 expression, proliferation information via Ki-67, and histopathology of the tumor. We used the following characterization based on available information shared to us from each biobank on the reporting of hormone status. HER2 was considered positive with staining intensity of 3+: Hormone receptor positive (i.e. estrogen receptor or progesterone receptor) (HR+) and HER2 positive (HER2+); HR+ and HER2 negative (HER2-); HR negative (HR-) and HER2+; and HR- and HER2-.

### Peptide Quantification and Mass Spectrometry Parameters

Prepared peptides were resuspended in 0.1% formic acid in water and quantified using a quantitative fluorometric peptide assay (Thermo Fisher, 23290). EvoTips were prepared as directed by the manufacturer with the exception that solvent volumes were doubled. Samples were analyzed on an Orbitrap Astral mass spectrometer (Thermo Fisher) coupled to an EvoSep One liquid chromatography (LC) system (EvoSep) running the whisper zoom 40 SPD with an Ion Opticks column (AUR3-15075C18-XT) set to 1,600 V and default parameters from the Label Free Quantification using Data Independent Acquisition (LFQ DIA) preset with 3 Th windows.

### Mass Spectrometry Data Analysis

Running in data-independent acquisition (DIA) mode, the Orbitrap Astral produces raw mass spectra from which peptide quantities can be computed based on a spectral library. Peptide quantities are then analyzed to produce gene group abundance data that are ultimately used for the classifier. The spectral library used herein was generated from an independent set of 280 patient samples consisting of individuals with breast cancer and healthy controls as previously described^13^. All peptide quantification and gene group abundance data were generated from DIA-NN (2.1.0) with default parameters except for the following settings: Maximum mass accuracy fixed to 11 and 10 ppm (MS1 and MS2 respectively), scan window of 7, up to one variable modification of methionine oxidation or N-terminal acetylation. A random subset of 100 patient samples used for spectral library generation was designated as “normalizing samples” for protein quantification. QuantUMS parameters were trained on these 100 samples and fixed for subsequent analysis^14^. The samples were each processed in independent DIA-NN sessions alongside all 100 normalizing samples (i.e. total of 101 samples) to obtain the MaxLFQ-normalized quantifications^15^.

The gene group-level abundance data from DIA-NN was preprocessed as follows. Non-missing abundances were log-transformed, and any remaining missing values were imputed using the minimum observed abundance for each gene-group minus one. Finally, the abundances were standardized by gene-group (zero mean and unit variance) to generate features suitable for machine learning classification models.

We developed QC metrics for both the plate level and the individual sample level. At the plate level, we evaluated mass spectrometry controls that included commercial enolase standards for retention time and total precursor ion intensity, commercial HeLa cell lysates were evaluated for peptide and protein IDs and selected peptides were assessed for consistent abundance values across the plates, and blank injections were added to compare total ion chromatograms (TIC) between blank and patient samples. Processing controls, commercial yeast lysates, and positive and negative controls were analyzed to ensure all sample processing steps were sufficiently completed. At the sample level, we required that each sample meet minimum peptide and protein IDs. Plates or samples not meeting these QC thresholds were re-ran. Two unsuccessful runs of the same sample resulted in discarding that sample.

### Reproducibility and Repeatability Analysis

For reproducibility experiments, two different operators ran the same samples from 8 donors (4 healthy and 4 breast cancer) over two different days. For repeatability experiments, the first operator ran samples from 4 donors (2 healthy and 2 breast cancer) in triplicate across three different plates, while a second operator ran a different set of samples from 4 donors (2 healthy and 2 breast cancer) in triplicate on a single plate. In total, 16 donors were run for reproducibility, and 24 donors were run for repeatability. Performance of each sample was assessed where each of the samples was classified as healthy or breast cancer. If the same sample was classified correctly across operators or days, then the sample was positive. If the samples were classified differently across operators or days, then the sample was negative.

### Training and Validation Split

We randomly assigned 845 of the samples to the training portion and the remaining 397 samples were assigned to the validation portion (a roughly 70:30 split), where their clinical information was blinded from sample preparation and analysis. Validation samples were batched separately from training samples to mimic real world situations around sample acquisition for an early detection test. Any samples that did not pass QC were removed from the analysis.

### Machine learning and cross-validation analysis

To avoid the potential of confounding by source, we implemented an Exponentiated Gradient method^16^, ensembling L2-norm regularized logistic regression, with the constraint of demographic parity across two sources. The default parameters in fairlearn library were used. To make predictions as part of cross-validation and to assess performance on the holdout validation, predictions of each component logistic regression were weighted according to the model weights estimated by the Exponentiated Gradient method. Source was treated as a sensitive feature.

We performed five-fold cross-validation of this model to assess the performance of our assay in distinguishing breast cancer from healthy samples in the training portion. At each iteration of the cross-validation, the data pre-processing described above was learned without the heldout fold, and the learned statistics were then applied to the heldout fold. The lambda parameter (strength of L2 regularization) was tuned by performing nested cross-validation (five folds) and selecting the parameter which maximized accuracy. A threshold for >90% specificity was determined using the outer-fold cross-validated predictions in healthy samples. The model was then trained on all of the training data, using the most frequently selected lambda from the previous step, and predictions were made on the validation samples. Validation samples were called positive if the predicted score was above the threshold determined from the training data and otherwise called negative. After these classifications were made, the validation data was unblinded, and sensitivity and specificity were calculated. All data analysis was performed using Python 3.12.4; the model was trained using the scikit-learn library (version 1.4.2) and the fairlearn library (version 0.13.0).

We assessed the robustness of our classifier by looking for associations between the cross-validated labels of the healthy samples using the heldout and potential confounders including source, race, age of the donor at time of collection, and batch. This confounder analysis is repeated in the validation set, using the predicted labels of healthy samples. We performed a chi-squared contingency table test for all these factors. The age of the donor was binarized based on the mean of the healthy samples.

### Gene Set Enrichment Analysis

Preranked gene set enrichment analysis (GSEA, version 4.4.0) was performed comparing all the healthy control samples to the breast cancer samples (including training and validation sets). Limma (version 3.64.1) of non-imputed data was used to perform differential expression of proteins (DEP) between the two groups, where Empirical Bayes moderation of the standard errors was applied with robust variance estimation and a trend prior (trend=TRUE, robust=TRUE). The ranked list was made using sign(logFC)*-log10(adj.P.Val) to rank the genes, and this ranked list was then used as input to preranked GSEA.

### Healthcare utility

To model the impact of the Astrin Biosciences’ breast cancer early detection test on finding dense breast cancers missed by the current screening guidelines, we simulated a population of 100,000 women with dense breasts screened via annual mammogram. Based on the literature, we simulated the dense breast population of women to have the following characteristics:

- 84% have BI-RADS density score 3 and 16% have density score 4^17^.
- 1.24% of BI-RADS density score 3 have incident breast cancer and 1.30% of BI-RADS density score 4 have incident breast cancer^17^.

Performance of diagnostics for dense breast cancer were derived from published literature to incorporate into the simulation:

- Sensitivity and specificity of screening mammogram in BI-RADS density score 3 are 69% (62%-76%) and 97% respectively; sensitivity and specificity in BI-RADS density score 4 are 47% (30%–65%) and 98%, respectively^17^.
- Sensitivity and specificity of MRI are 97% (86%-99%) and 69% (46%-85%), respectively^18^.
- Sensitivity and specificity of CEM are 91% (77%-97%) and 74% (52%-89%), respectively^18^.
- Sensitivity and specificity of ABUS are 67% (53%-79%) and 89.9% (89.1%-90.6%), respectively^19^.

Using the above as well as the sensitivity and specificity of our test as measured in the validation study, we simulated the number of false negative breast cancers and false positives in women with dense breasts. In this population, we assume all women receive a mammogram, and then following a negative mammogram, women may receive one of four different supplemental screens, with adherences ranging from 1% to 100%. This simulation is repeated 100,000 times, and we summarize the number of false negatives and false positives by median, 2.5 percentile, and 97.5 percentile. Uncertainty of sensitivity and specificity for all diagnostics are incorporated into the simulation.

### Study approval

Samples were obtained from commercial biobank locations who consent patients under appropriate institution review board approved protocols. All the patient data was provided to Astrin Biosciences in a de-identified manner.

### Data Availability

The data sets analyzed in this study are available upon reasonable request by email to the corresponding author.

## Results

### Development of a high throughput pipeline for protein-based detection from plasma

For MS-based proteomics to be considered a viable platform for early cancer screening, it was necessary to develop and rigorously validate a high throughput assay capable of analyzing hundreds of patient samples per week. To do this, we utilized an automated robotics platform that allows up to 76 samples to be processed in parallel (Fig. 1A, B, Supplemental Fig. 1). To validate that sample preparation and analysis meet strict criteria around repeatability (under same conditions) and reproducibility (under different conditions), multiple controls were included. Utilizing commercial digests of yeast enolase and HeLa lysates, we can measure the quantification of specific peptides and show that the retention time stability across all of our sample batches is consistent (Supplemental Fig. 2A-F). To confirm that we are minimizing sample carryover from run to run on the MS, we analyzed the TIC of blank injections and patient samples (healthy and cancer). The blank injections were <1% of the total TIC signal from patient samples (Supplemental Fig. 2G).

**Figure 1.**
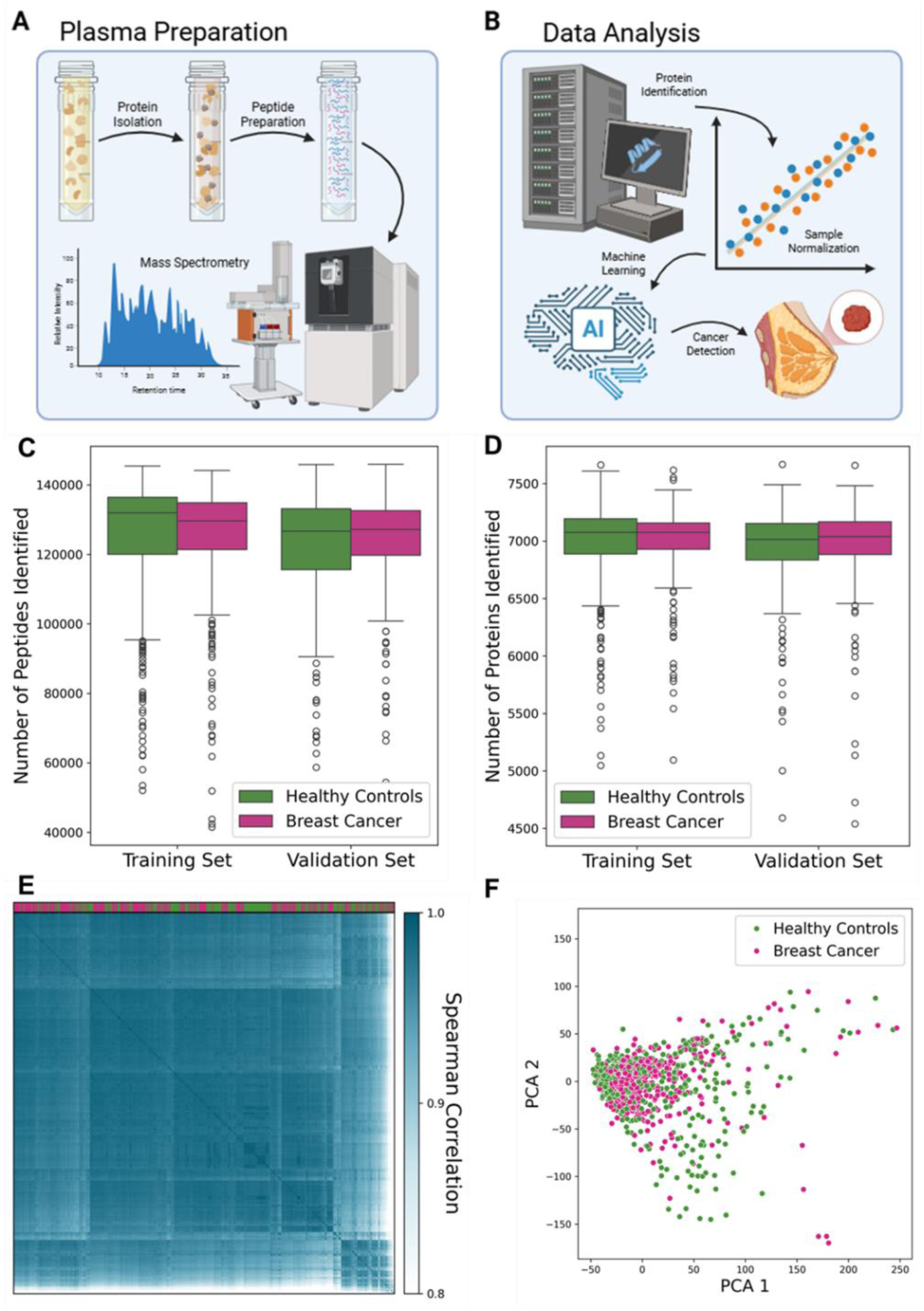
Overview of the sample processing methodology (A) after receiving the banked plasma samples through injection onto the mass spectrometer followed by data processing, normalization and then cancer prediction (B). Created using BioRender. The number of peptide (C) and proteins (D) across health and breast cancer samples (both training and validation cohorts) were nearly identical. Spearman correlation (E) and principal component analysis (PCA, F) were assessed for any discrepancies across the samples which again were nearly identical.

While this demonstrates that the Evosep One liquid chromatography (LC) system and the Orbitrap Astral mass spectrometer (MS) are well suited to the challenging conditions required to meet the high standards required for a clinical assay, sample preparation remained as the most likely source for variation. To circumvent this, we prepared and included a known concentration of intact yeast protein extract that was spiked onto each plate and processed in parallel with the patient samples. This allowed us to track sample recovery, reduction and alkylation efficiency, peptide recovery, and protein and peptide identifications (Supplemental Fig. 3A). In addition, positive and negative controls made from pools of 9 late-stage breast cancer samples (stage III and IV) and healthy patient samples, respectively, were analyzed. These control pools provided information on the consistency of the preparation process over time including peptide and protein IDs (Supplemental Fig. 3B, C). These samples were not included as part of the training data, but we verified that they classified correctly once the model training was completed (Supplemental Fig. 3D).

This laboratory developed assay was validated in accordance with the Clinical Laboratory Improvement Amendments (CLIA) standards for high-complexity testing, and the assay’s analytical performance has been characterized and documented. Indeed, repeatability and reproducibility analysis showed full agreement between repeated measurements of the same samples. The n=8 samples measured in triplicate showed an average positive agreement (APA) of 100% (4/4), and an average negative agreement (ANA) of 100% (4/4). The samples measured by two different operators also had APA of 100% (4/4) and ANA of 100% (4/4). Qualitatively, model scores showed strong agreement within donors whether from the same operator or across operators (Supplemental Fig. 4).

Using this high throughput proteomic pipeline, we trained on plasma samples that passed quality control (QC) metrics for peptide and protein counts (Fig. 1C, D). This results in a total of 845 women, including 379 newly diagnosed, treatment naïve breast cancer patients (pathological stage 0: n = 45, I: n = 130, II: n = 152, III: n = 35, IV: n = 14) and 466 healthy controls. The patient demographics are summarized in Table 1. The plasma samples were de-identified, randomized, and blinded before processing and running on the MS. All MS files were run sequentially and normalized to identify and quantify each individual patient’s proteome.

**Table 1.** Patient demographics and splits used in the training and validation sets.

| Demographics |  | Training Set |  | Validation Set |  |
| --- | --- | --- | --- | --- | --- |
|  |  | Healthy | Cancer | Healthy | Cancer |
| **Sample size** | Count, *n* | 466 | 379 | 195 | 202 |
| **Age** | Mean (SD) | 55.4 (10.8) | 57.2 (12.4) | 56.4 (8.8) | 59.8 (13.0) |
| **BMI** | Mean (SD) | 26.2 (4.4) | 27.8 (6.3) | 26.2 (4.3) | 26.7 (5.4) |
| **pStage** | 0, *n* (%) |  | 45 (11.9%) |  | 13 (6.4%) |
|  | I, *n* (%) |  | 130 (34.3%) |  | 69 (34.2%) |
|  | II, *n* (%) |  | 152 (40.1%) |  | 98 (48.5%) |
|  | III, *n* (%) |  | 35 (9.2%) |  | 16 (7.9%) |
|  | IV, *n* (%) |  | 14 (3.7%) |  | 2 (1.0%) |
|  | Unknown, *n* (%) |  | 3 (0.8%) |  | 4 (2.0%) |
| **Pathology** | LCIS, *n* (%) |  | 1 (0.3%) |  | 1 (0.5%) |
|  | DCIS, *n* (%) |  | 38 (10.0%) |  | 10 (5.0%) |
|  | ILC, *n*(%) |  | 33 (8.7%) |  | 20 (9.9%) |
|  | IDC, *n*(%) |  | 270 (71.3%) |  | 153 (75.7%) |
|  | Mixed, *n*(%) |  | 4 (1.0%) |  | 1 (0.5%) |
|  | Other / Unknown, *n*(%) |  | 33 (8.7%) |  | 17 (8.4%) |
| **Subtype** | HR+/HER2- |  | 190 (50.1%) |  | 121 (59.9%) |
|  | HR-/HER2- |  | 35 (9.2%) |  | 16 (7.9%) |
|  | HR+/HER2+ |  | 27 (7.1%) |  | 6 (3.0%) |
|  | HR-/HER2+ |  | 19 (5.0%) |  | 3 (1.5%) |
|  | Unknown |  | 108 (28.6%) |  | 56 (27.7%) |
| **Race** | White, *n* (%) | 429 (92.1%) | 355 (93.6%) | 176 (90.3%) | 197 (97.5%) |
|  | Black/African American, *n* (%) | 12 (2.6%) | 8 (2.1%) | 4 (2.1%) | 1 (0.5%) |
|  | Asian, *n* (%) | 21 (4.5%) | 15 (4.0%) | 10 (5.1%) | 4 (2.0%) |
|  | Other/Unknown, *n* (%) | 4 (0.8%) | 1 (0.3%) | 5 (2.5%) | 0 (0.0%) |
| **Source** | ProteoGenex, *n* (%) | 316 (67.8%) | 240 (63.3%) | 105 (53.8%) | 129 (63.9%) |
|  | BioIVT, *n* (%) | 150 (32.2%) | 139 (36.7%) | 90 (46.2%) | 73 (36.1%) |

For all patient samples that were ran, the median (Q1-Q3) number of proteins detected between breast cancer and healthy samples were 7,064 (6,904-7,160) and 7,054 (6,876-7,187) proteins, respectively, with 9,266 unique proteins observed across both cohorts. Most patients had more than 6,500 proteins detected per sample (92.5%, 1149/1242) with no significant differences in means across the two cohorts (Welch’s t-test p-value: 0.81 (training), 0.50 (validation)), demonstrating both the consistency of our sample preparation and the sensitivity of our MS method (Fig. 1C, D). Semi-quantitative assessment of protein intensities showed that we captured a dynamic range of protein abundances spanning over 8 orders of magnitude (Supplemental Fig. 5), highlighting the workflow’s potential to capture low abundance protein fractions. Spearman correlation (Fig. 1E) and principal component analysis of all proteins in the proteome (PCA, Fig. 1F) resulted in no discernible differences between the healthy and breast cancer samples validating the quality of sample and batch preparation in this cohort.

### Model training and held-out validation for breast cancer prediction

In cross-validation of the training samples, we observed an AUC of 0.961 (95% Bootstrap CI: 0.947-0.97) (Fig. 2A) in distinguishing breast cancer from healthy samples. A receiver operating curve (ROC) showed strong discriminative performance, demonstrating the model’s accuracy for breast cancer prediction (Fig. 2A). Across all patient samples in the training cohort, the model achieved a sensitivity of 89.2% (95% Wilson CI: 85.7-91.9%) with a specificity of 90.1% (95% Wilson CI: 87.1-92.5%) (Fig. 2B).

**Figure 2.**
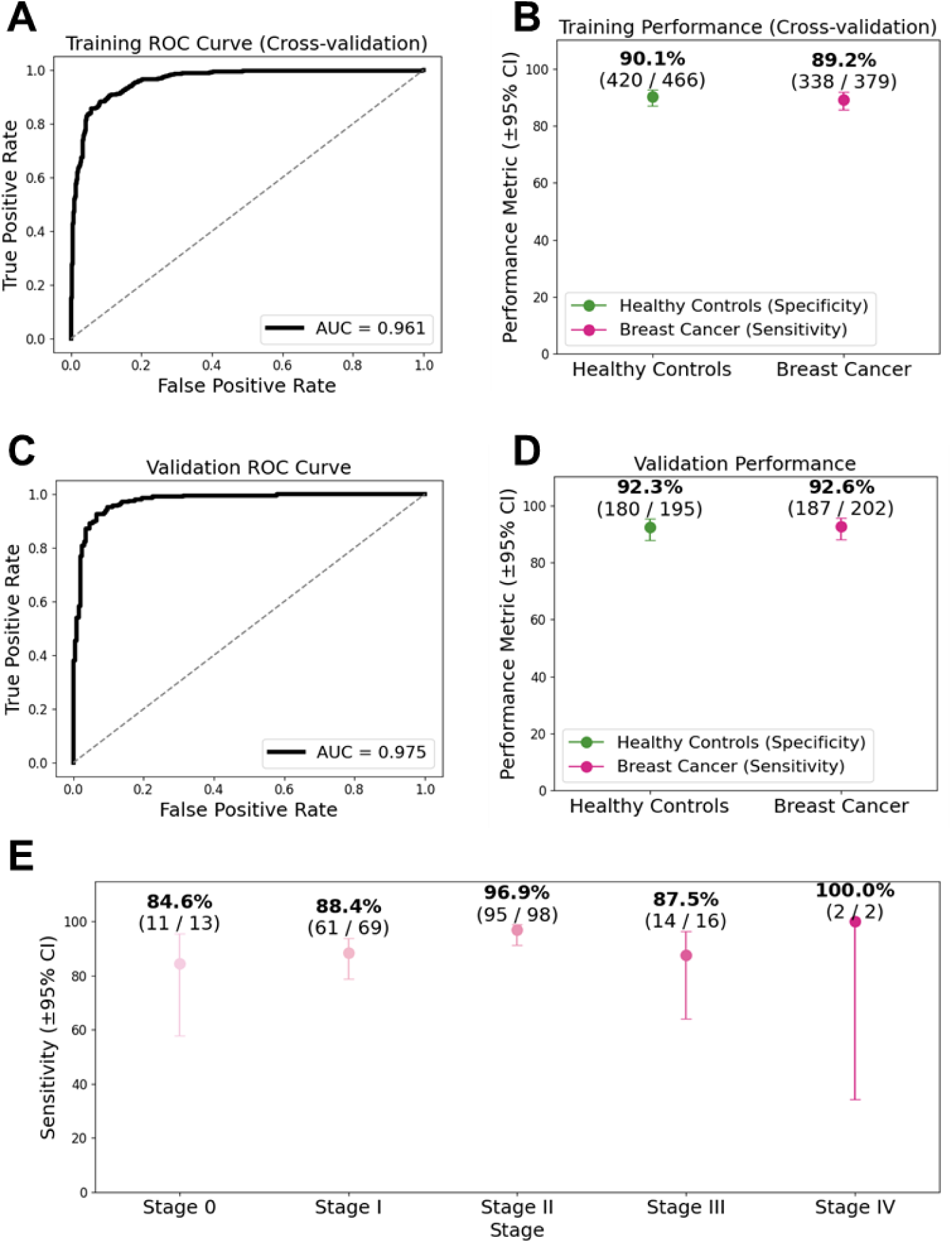
Training and validation performance. Receiver operator curve (ROC) was assessed for the training (A) and validation (C) with performance of sensitivity and specificity for the training (B) and validation samples (D). Sensitivity remained high across all breast cancer stages (E).

To validate our training model, we analyzed 397 samples consisting of 195 healthy and 202 breast cancer patients (pathological stage 0: 13, stage I: 69, stage II: 98, stage III:16, and stage IV: 2; stage unknown: 4) as a held-out validation set. Overall performance was 92.6% sensitivity at 92.3% specificity with an AUC of 0.975 (95% Bootstrap CI: 0.961-0.987) and a negative predictive value (NPV) of 99.96% (95% CI: 99.93-99.98) (Fig. 2C, D). When broken down by stage, we observed no discernible differences of performance accuracy across stages, which may relate to overall tumor burden (stage 0-IV sensitivities: 84.6%, 88.4%, 96.9%, 87.5%, and 100.0%, respectively) (Fig. 2E). Performance across breast cancer molecular (HR+/HER2+, HR+/HER2-, HR-/HER2+, and HR-/HER2-) and pathological (LCIS, DCIS, ILC and IDC) subtypes were also similar between the different groups (Fig. 3A, B). Potential confounders were also assessed in our training and validation cohorts including age, source, race, and plate (or batch) (Fig. 4A, B and Supplemental Figure 6A, B). We observed no difference between the predicted label and the tested variables among healthy samples in either the training or validation (Table 2).

**Figure 3.**
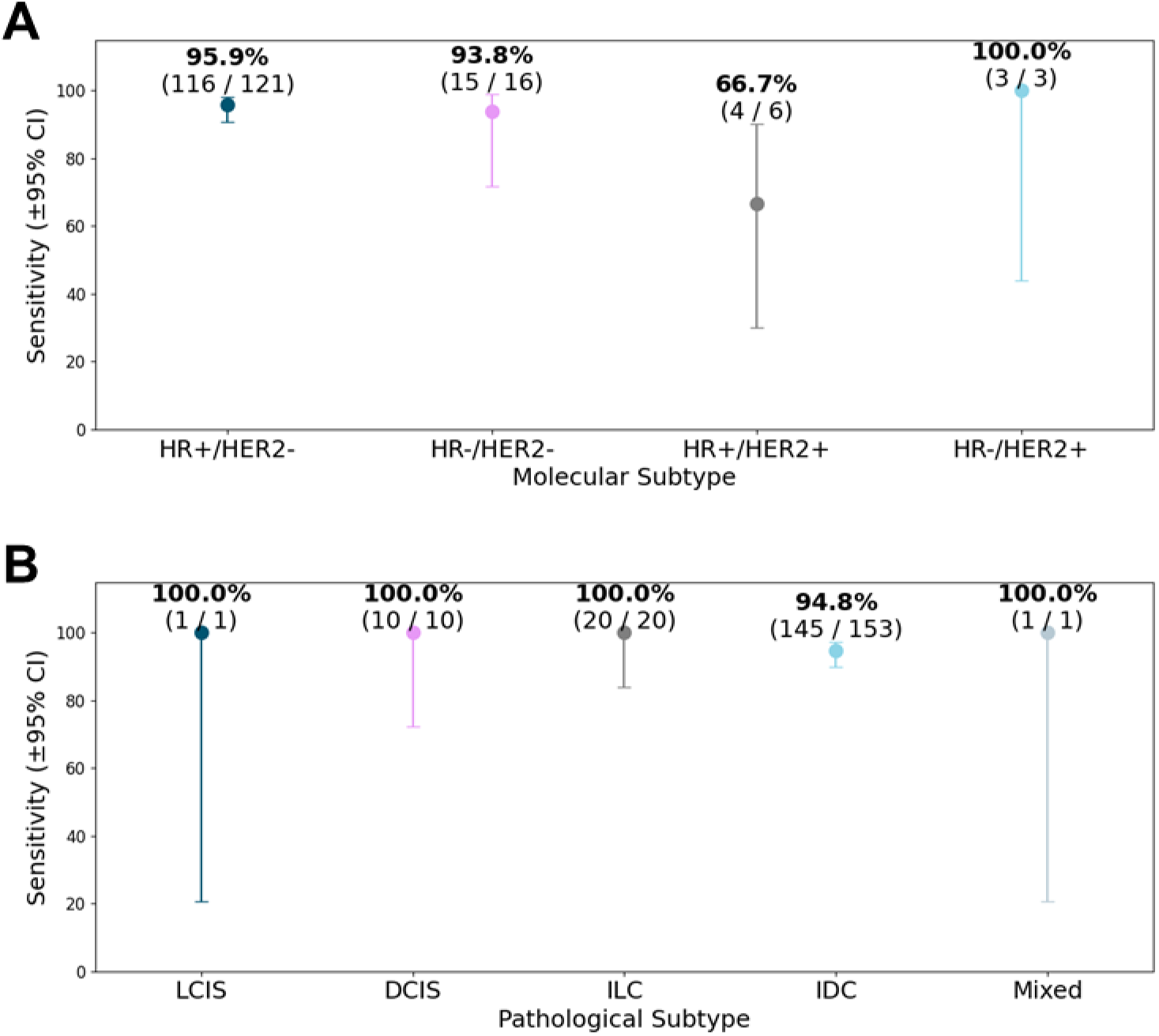
Sensitivity across breast cancer subtypes. Sensitivity analyses of breast cancer molecular (A) and pathological (B) subtypes show no discernable differences among the different groups.

**Figure 4.**
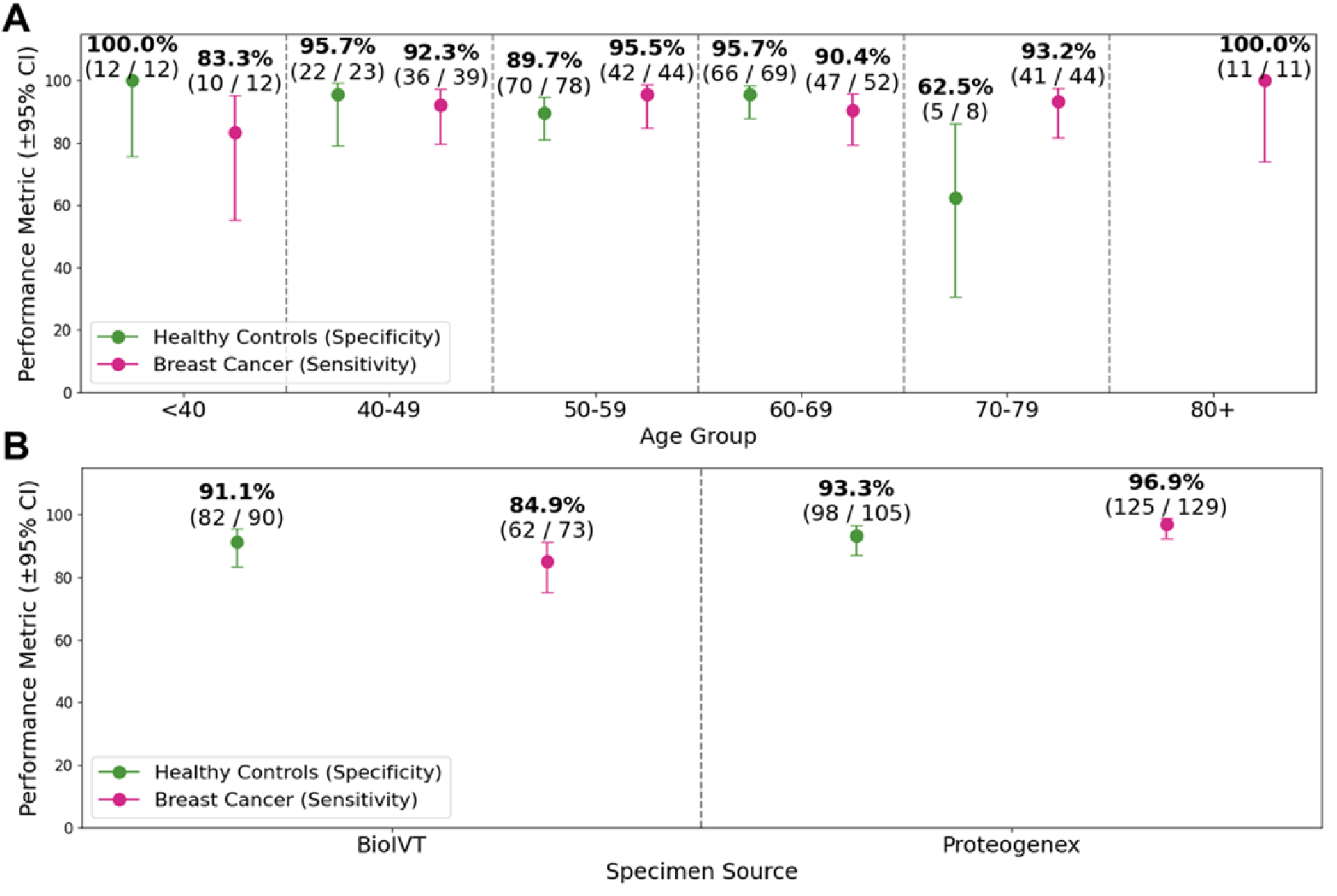
Assessment of confounders based on age and sample source. To ensure that the training model was not influenced by confounders, we assessed model performance by age (A) and sample source (B).

**Table 2.** Confounder analysis.

| Confounder | Training Set | Validation Set |
| --- | --- | --- |
| Age of donor | 0.484 | 1.000 |
| Plate | 0.629 | 0.282 |
| Race | 0.940 | 0.394 |
| Source | 0.150 | 0.721 |

### Pathway analyses of breast cancer samples

To ensure we are identifying and enriching cancer-associated proteins and pathways, we performed gene set enrichment analyses (GSEA) of the breast cancer samples. Based on 50 Hallmark pathways, we first assessed if the number of proteins detected met the minimum recommended threshold of 15 genes for each of the pathways (Fig. 5A). While this threshold was met for almost all pathways, on average ∼68% of the genes in each pathway were represented. GSEA identified enriched cancer-associated pathways including epithelial-to-mesenchymal transition (EMT), PI3K-AKt signaling, KRAS up signaling, and WNT/beta-catenin signaling (Fig 5B-D). Importantly, several immune-activation or regulatory pathways (Interferon gamma, alpha, TGF beta) were not significantly enriched in breast cancer samples, or even decreased in these plasma samples (IL6, IL2, TNFalpha signaling) (Supplemental Fig. 7A-C). Altogether, the plasma proteins captured were associated with known breast cancer pathways with limited or even negative associations with immune cell activity.

**Figure 5.**
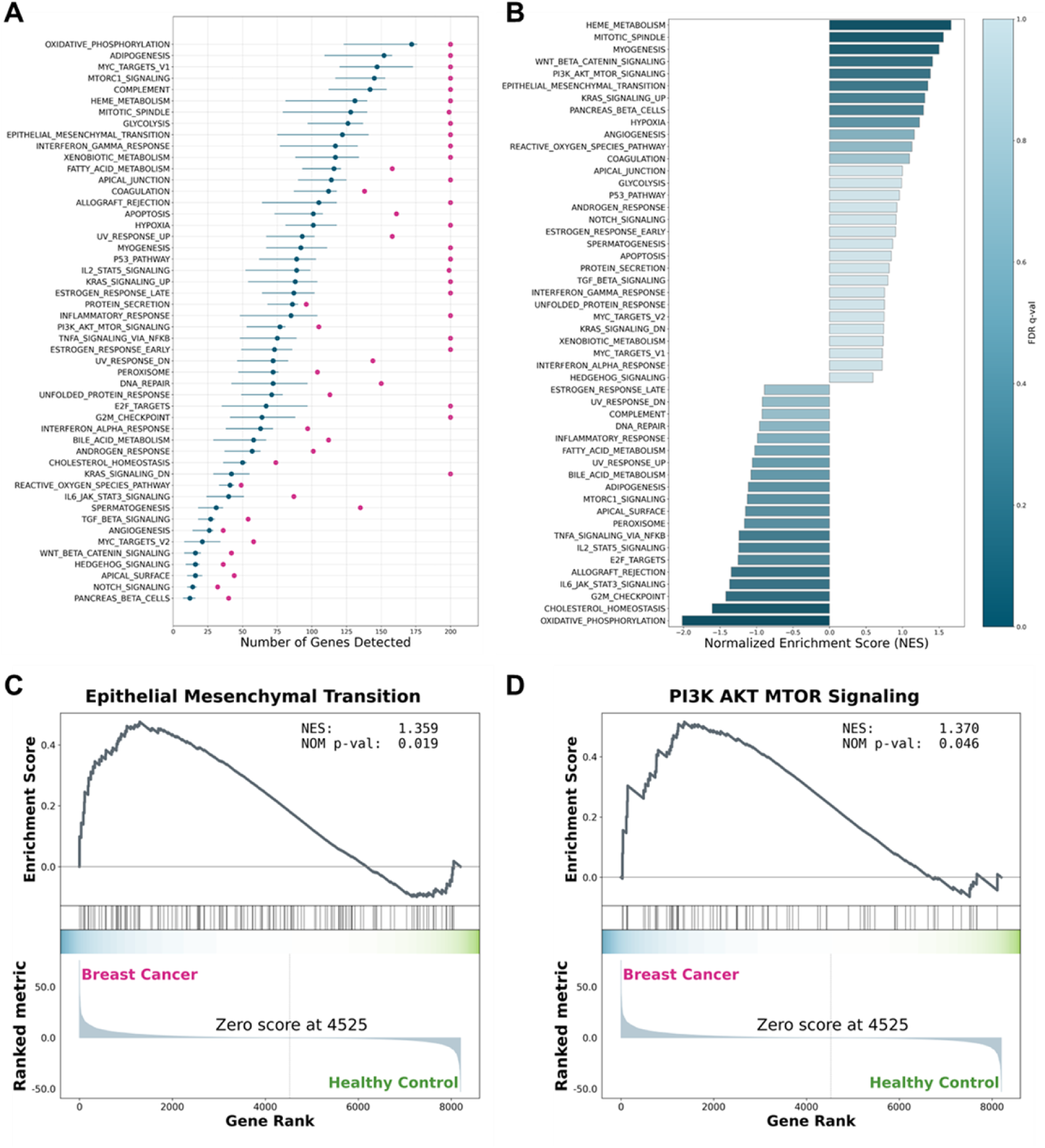
Pathway analysis of breast cancer enriched proteins. Gene set enrichment analysis was performed on all pathways with >15 genes (A) and found enrichment of pathways including epithelial-to-mesenchymal transition (EMT) (B, C) and PI3K-AKT signaling (B, D).

### Healthcare utility of Astrin Biosciences’ breast cancer early detection test

While breast density was lacking in our particular cohort to assess model performance, we have published in an earlier manuscript that the performance of our test is independent of breast density^13^. Based on that information and our sensitivity and specificity, we modeled the utility of our test to detect breast cancers in women with dense breasts in a screening population. We observed a dramatic increase in the proportion of dense breast cancers detected when incorporating our test as a supplemental screening test following a negative mammogram (Fig. 6A). In our analysis, we found that without supplemental screening, 431 (95% CI: 347-521) breast cancers were misdiagnosed. Increasing uptake in all pathways resulted in fewer missed breast cancers, but with ABUS, 142 (95% CI: 87-212) breast cancers were still missed at 100% uptake. Conversely, while MRI and CEM missed only 13 (95% CI: 2-49) and 39 (95% CI: 11-105) breast cancers respectively, there were 32,526 (95% CI: 16,958-54,361) and 27,724 (95% CI: 13,701-49,766) false positives at 100% uptake (Fig. 6A, B). Meanwhile, false positives remain low with our deep proteome analysis (4,898; 95% CI: 3,582-7,333) at 100% uptake with a similarly low number of breast cancers (49; 95% CI: 25-89) being misdiagnosed. Even with a moderate uptake (40%) of our test, we would reduce the number of misdiagnosed dense breast cancers detected to 278 (95% CI: 221-342) (Fig. 6B).

**Figure 6.**
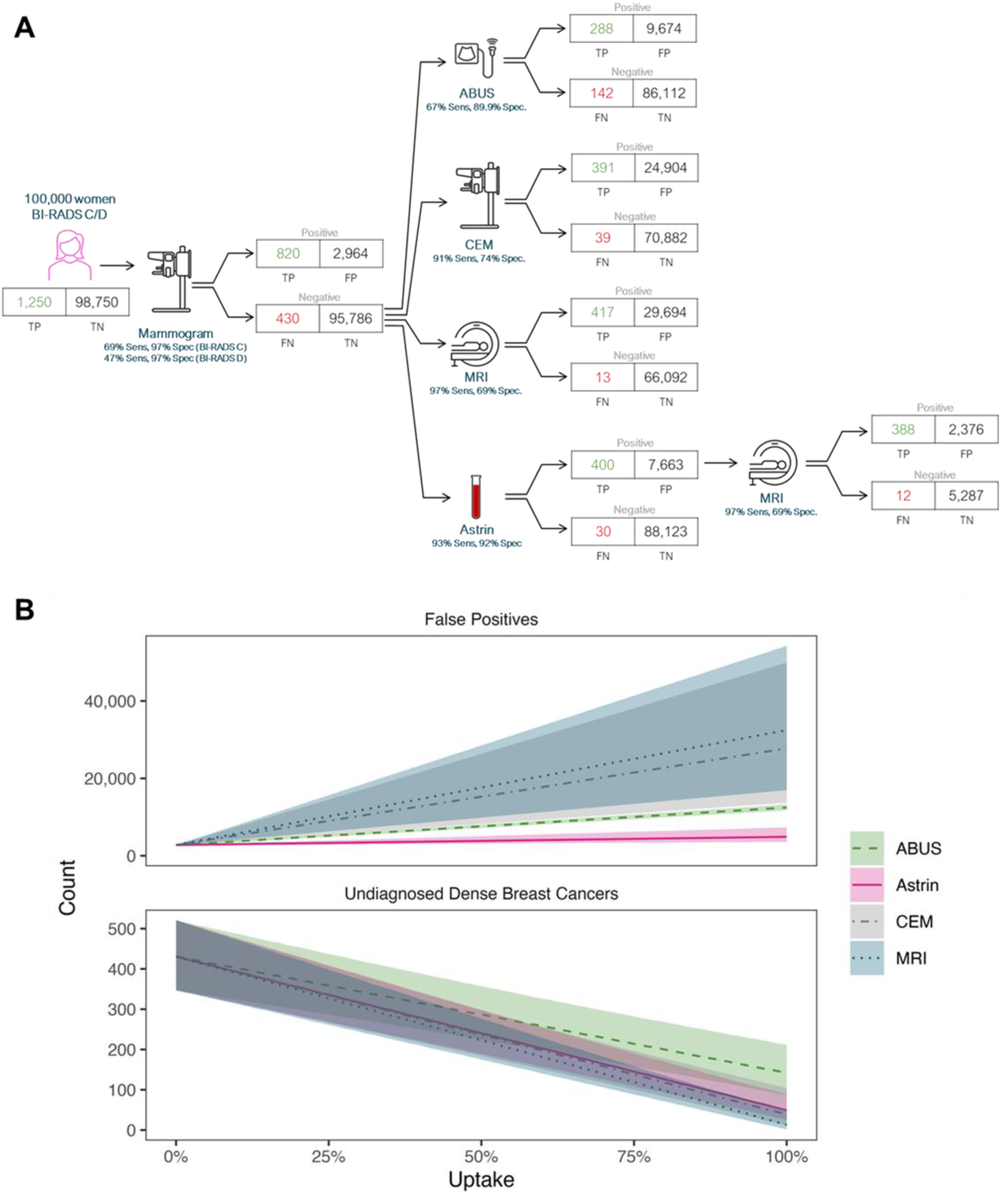
Healthcare utility of deep proteome based breast cancer supplemental screening test. Women with dense breasts were assumed to receive standard of care with low uptake of supplemental screening or receive our test following a negative mammogram (A) and found to have a much higher rate of detected dense breast cancers with low rates of false positives when screened via Astrin (B).

## Discussion

We have developed and built a reproducible and repeatable protein-based liquid biopsy assay that is robust for breast cancer detection, especially at early stages including pathological stages 0, I, and II. There were also no discernable differences in performance across molecular or pathological subtypes. Our machine learning classifier was able to discern cancer-specific signals and was not confounded by other variables including age, sample source, and batch. This assay is automated and capable of analyzing several hundred samples in parallel. Holdout validation of this test maintained high sensitivity and specificity, paving the way for such a test to be offered as a supplemental screening option for women with a negative mammogram with moderate to high risk of harboring breast cancer. This is especially important for women with dense breast tissue and an invasive lobular carcinoma (ILC) pathology, which are extremely difficult to detect via mammogram due to their distinct biology and growth pattern as compared to intraductal carcinoma (IDC)^20^. Importantly, our test identified 100% of the ILC cases and we plan to enrich for this histology in future prospective studies to confirm performance as it will be critical to have an early detection test sensitive enough to detect ILCs.

For over four decades, mammograms have been used to screen for breast cancer, enabling early diagnosis in many individuals, thereby increasing breast cancer survival. While mammogram sensitivity in women with non-dense breast tissue is high, women with dense breast tissue are at risk of missed tumors as mammograms have been shown to miss more than half of diagnosed cancers (8.4/1000 for mammogram and 17.4/1000 AB-MRIs per 1000 supplemental exams)^6^. This underscores the considerable opportunity to enhance cancer detection in women with dense breast tissue. Currently, women with dense breasts are referred to an additional screening modality, typically ultrasound, which increases the number of cancers detected but still with low sensitivity. Although supplemental imaging techniques such as automated whole breast ultrasound, CEM, and abbreviated MRI (AB-MRI) each offer incremental gains, they are constrained by important trade-offs including operator dependence, cost, higher recall burdens, limited accessibility, and reliance on contrast agents. These persistent challenges highlight the need for innovative approaches capable of improving detection which are convenient, affordable, specific, and do not strain clinical workflows. In this context, our proposed breast cancer early detection test offers a promising path forward with a high sensitivity in early-stage breast cancers, including women with dense breasts, while maintaining high specificity and NPV. As a liquid biopsy test, implementation of the plasma derived deep proteome analysis into the clinic would be straightforward and could be widely implemented after mammogram and before MRI as a supplemental screening test. For example, a dense breast patient who needs supplemental screening would receive our test following a negative mammogram with the goal of reducing unnecessary MRIs. In the case of a positive result, the patient would then be referred to an MRI for tumor identification and biopsy.

In recent years, many blood-based cancer screening tests have entered the market. Shield^TM^ by Guardant specifically detects the presence of colorectal cancer while FirstLook by DELFI Diagnostics detects lung cancer. Notably, Grail Galleri® and Exact Sciences CancerGuard^TM^ are both multi-cancer early detection (MCED) tests with varying sensitivities based on the cancer type evaluated and cancer stage. All of these tests detect cancer via cfDNA, such as fragmentomics, methylation, or somatic alteration as the primary analyte, with some supplemented by a panel of blood-based proteins, such as in the CancerGuard^TM^ test. Importantly, it should be emphasized that the sensitivity for early-stage breast cancer detection is low for both Galleri® and CancerGuard^TM^ implying that cfDNA-based assays, even with a small sub-sampling of proteins, may not be sufficient as the primary analyte for early-stage breast cancer detection, likely due to the low shedding rate of cfDNA in early-stage cancers^21^. Indeed, the sensitivity for early-stage breast cancer detection using Galleri® is 2.6%-47.5% for stage I and II patients, respectively^9^ and for CancerGuard^TM^ is 8.3%-40.0% for stage I and II patients, respectively. Further, the MCED tests require several milliliters (mL) to several tubes of blood. The described breast cancer early detection test has the unique advantage of finding early-stage breast cancer with <1 mL of blood with sensitivities over 90% across stages 0-II, and will be comparable in cost to MRI or other screening tests.

There are a few notable limitations to this study. First, the analysis is conducted on samples purchased from biobanks. While we ruled out confounding with available clinical information, we cannot rule out the possibility that our classifier is confounded by other unmeasured factors, such as selection bias and variations in collection and storage techniques. Second, this analysis has been conducted on women where BI-RADS density was not recorded. Importantly, our analysis of a separate study showed that it is likely that our test performance is independent of breast density^13^. Future studies will be required to confirm performance and suitability for supplemental analysis of dense breast cancer patients at the same time. Third, for validation, we randomly allocated our dataset into training and validation. These validation samples were batched separately from the training samples and so represent generalization to future batches. However, future validations will be required to confirm generalizability to future groups of individuals from those observed in training using external datasets. Fourth, the racial demographics of healthy and breast cancer patient training and validation sets were predominantly white (92.8% and 93.2%, respectively), potentially limiting the validity of these findings for minority groups. Future efforts will be aimed at prospective collection of samples that increase source variety with more complete clinical annotations and reducing racial disparities in our validation cohorts. Finally, our current methodology requires blood to be drawn into EDTA tubes, centrifuged, and frozen the same day. Future work will also evaluate other ways to mitigate protein variations that come from blood stored in EDTA tubes, with the goal of increasing the accessibility and consistency of recruited samples.

## Conclusions

We describe a novel deep proteome-based breast cancer early detection test that can serve as a supplemental screening test particularly in patients with dense breasts (BIRADS C,D) as seen on screening mammography. The test has a high sensitivity and high specificity with an NPV of 99.7% with the potential to reduce a significant number of unnecessary MRIs and biopsies.

## Authors’ Contributions

Conceptualization: **AH**, **JMD**; Data curation: **AH**, **YT**, **JC, KM**, **KJK**, **NRB, AG, BRK**; Formal analysis: **AH**, **YT**, **KM**, **NRB, EB**; Supervision: **CGG, JH, JMD**; Validation: **YT, JH, JMD**; Investigation: **AH**, **GS**, **CR, JMD**; Visualization: **YT**, **JC, EB, KJK**, **JMD**; Methodology: **AH**, **GS**, **CR, JMD**; Writing – original draft: **AH**, **YT**, **JC, KJK**, **HB**, **JMD**; Project administration: **KJK**, **CGG**, **ARH**, **JMD**; Writing – review and editing: **AH**, **YT**, **JC, EB, KM**, **GS**, **CR, KJK**, **NRB**, **CGG**, **AG, HB**, **ARH**, **PA, JH**, **BRK**, **JMD**.

## Acknowledgements

We would like to thank all the brave patients who donated blood for this study. We would also like to thank the entire staff at Astrin Biosciences for their continued efforts to end cancer as we know it.

---

**Supplemental Figure 1.**
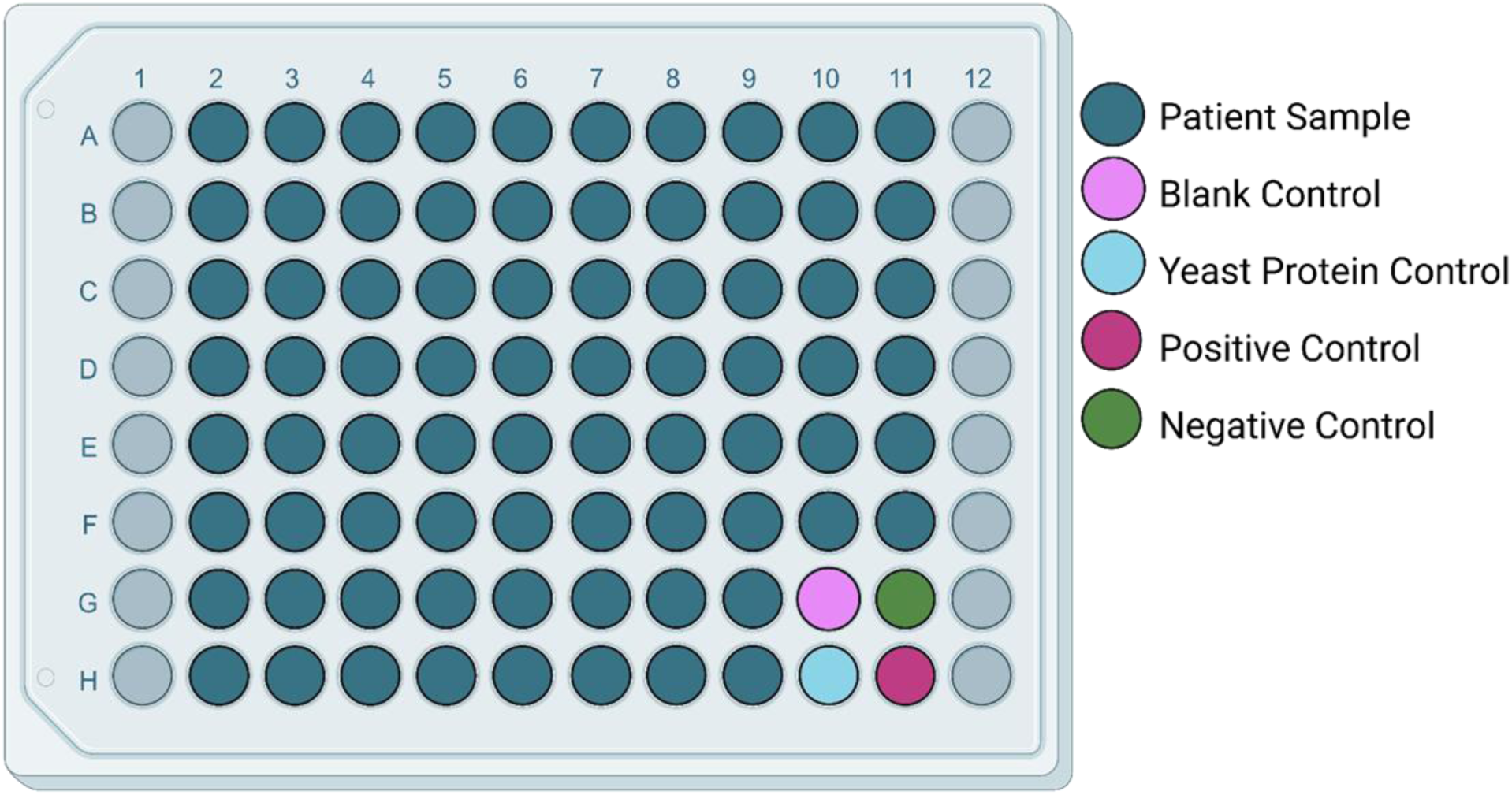
Plate layout highlights the locations of the controls and patient samples. Columns 1 and 12 are reserved for standard curves during peptide normalization.

**Supplemental Figure 2.**
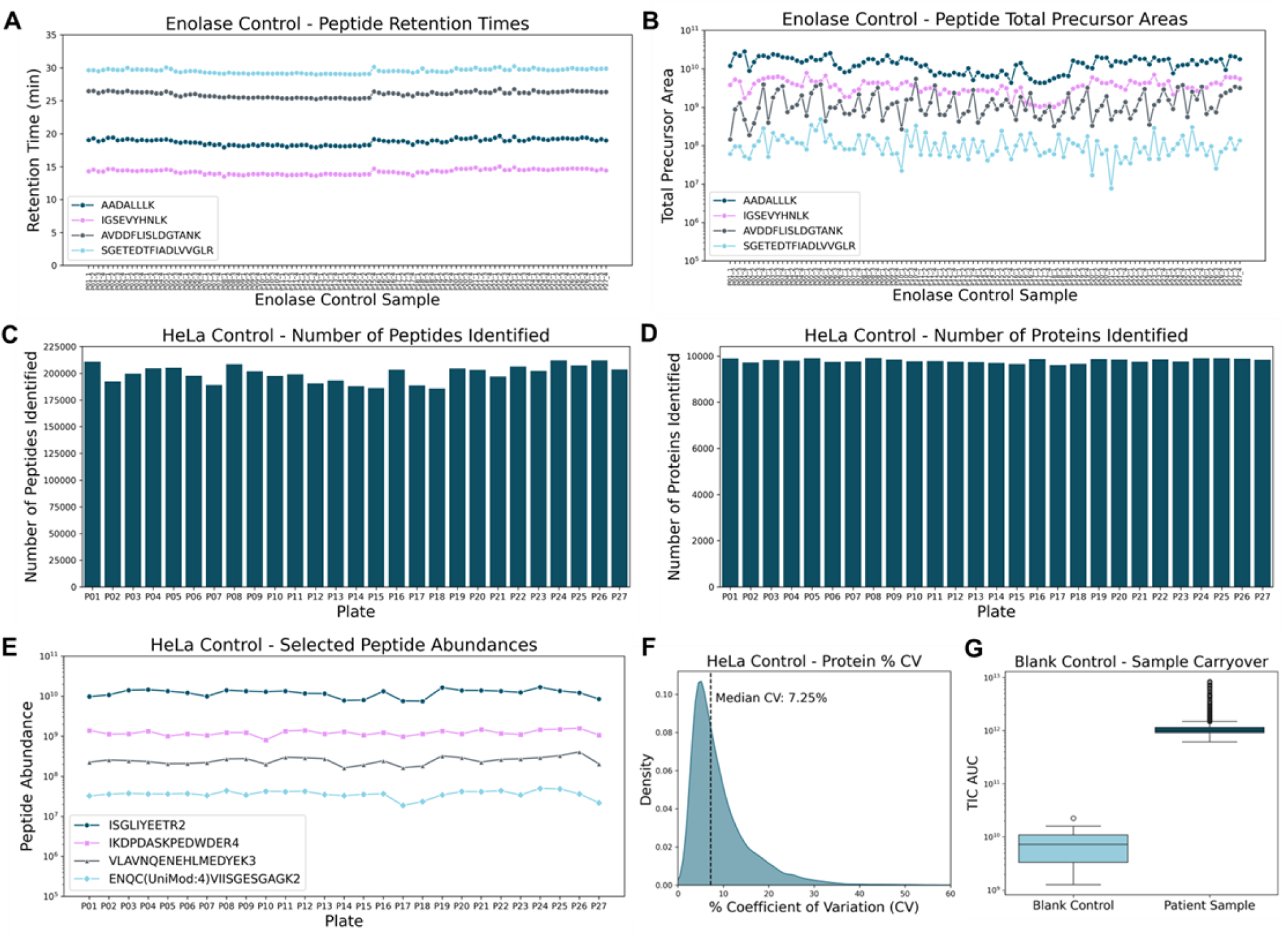
Plate level controls to assess mass spectrometry functionality. Commercial enolase standards were assessed for retention time (RT) differences (A) and total precursor areas for four selected peptides (B). Commercial HeLa cell lysates were assessed for the total number of peptides (C), proteins (D), and peptide abundances of four selected peptides (E). The percent coefficient of variation (%CV) (F) was also evaluated on HeLa cell lysates to measure the consistency and reproducibility of the mass spectrometry runs. Total ion chromatograms (TIC) were compared between blank injections and patient sample injections to determine possible carryover of protein signals. The blank samples represent <1% of the patient samples TIC (G).

**Supplemental Figure 3.**
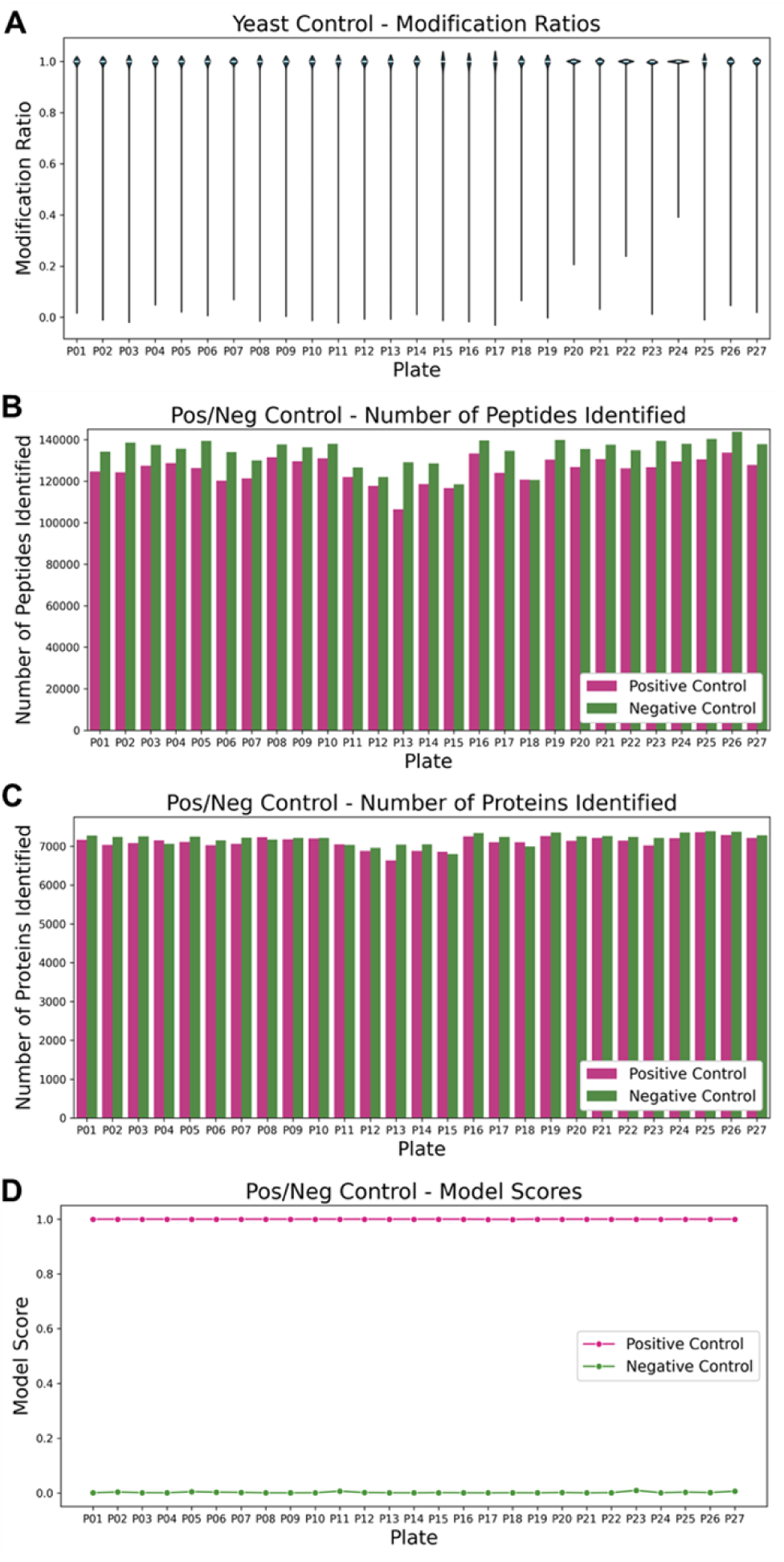
Sample level controls to assess processing functionality. Commercial yeast lysate was added to an individual well to mimic sample preparation. Cysteine modifications were evaluated to measure proper alkylation and reduction efficiency (A). Positive (B) and negative (C) control samples consisting of breast cancer and healthy patients, respectively, were evaluated for the number of peptides (B) and proteins (C) between both groups. The performance of the controls were also evaluated (D).

**Supplemental Figure 4.**
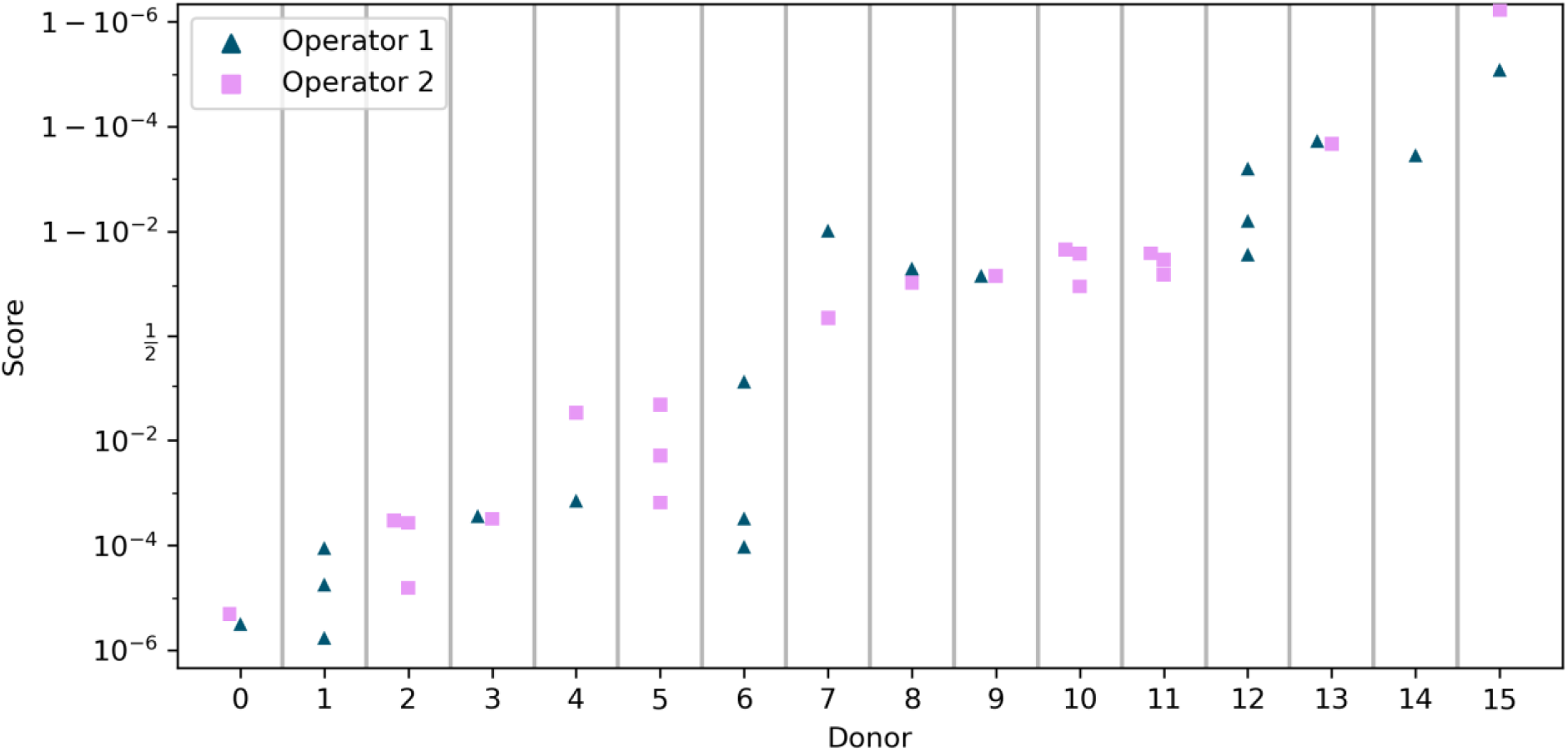
Repeatability and reproducibility samples were processed in triplicate by a single operator (n=8) or reproduced by two operators (n=8). The model scores show tight clustering within a donor, indicating that most of the variability can be assigned to sample variability rather than operator or test variability. After classification using the model threshold, 100% of the donors showed full agreement between the repeated measurements.

**Supplemental Figure 5.**
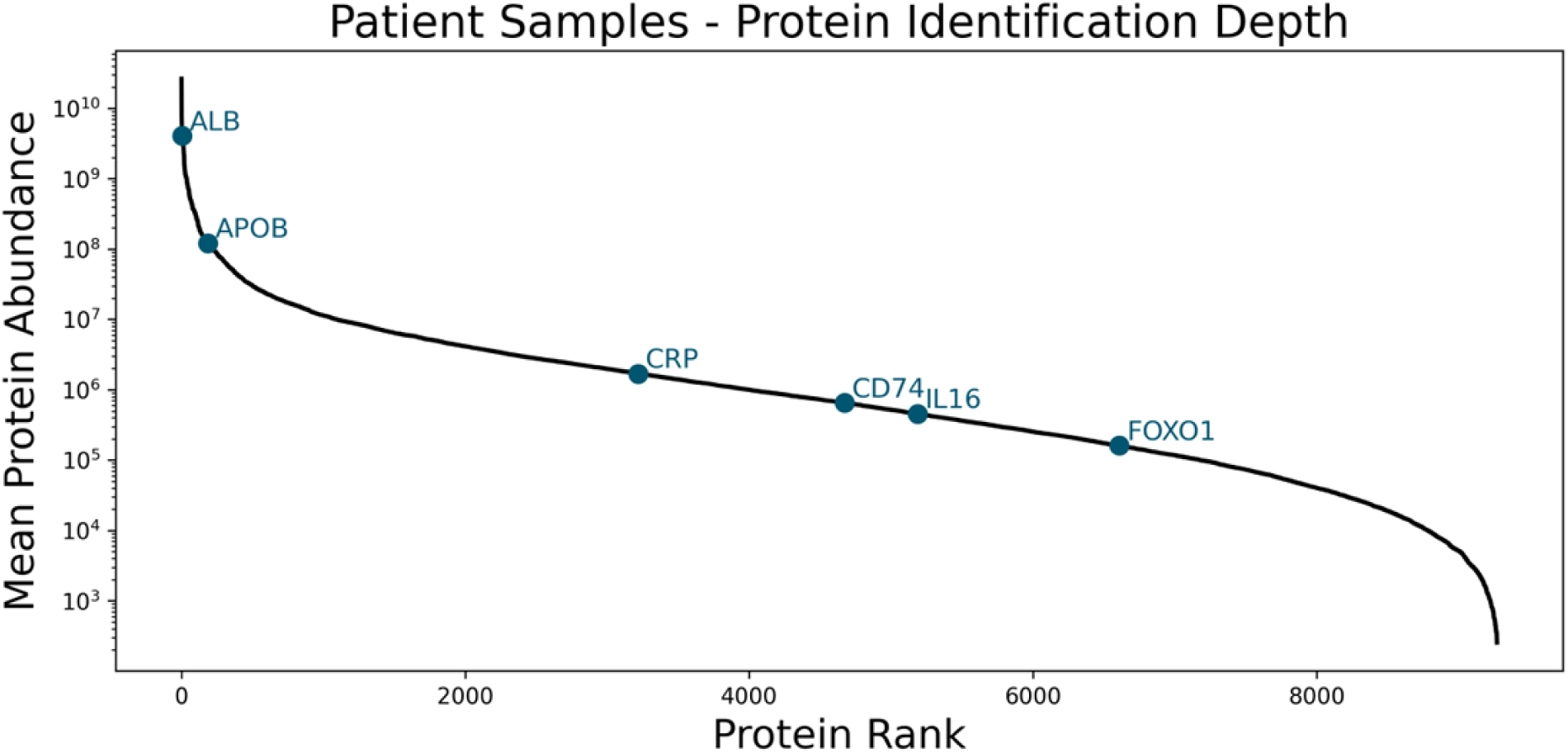
Depth of proteome coverage. The S-plot shows protein abundance over 8 orders of magnitude over 9,000 proteins. Typical proteins described in the Human Protein Atlas are shown and labeled in blue.

**Supplemental Figure 6.**
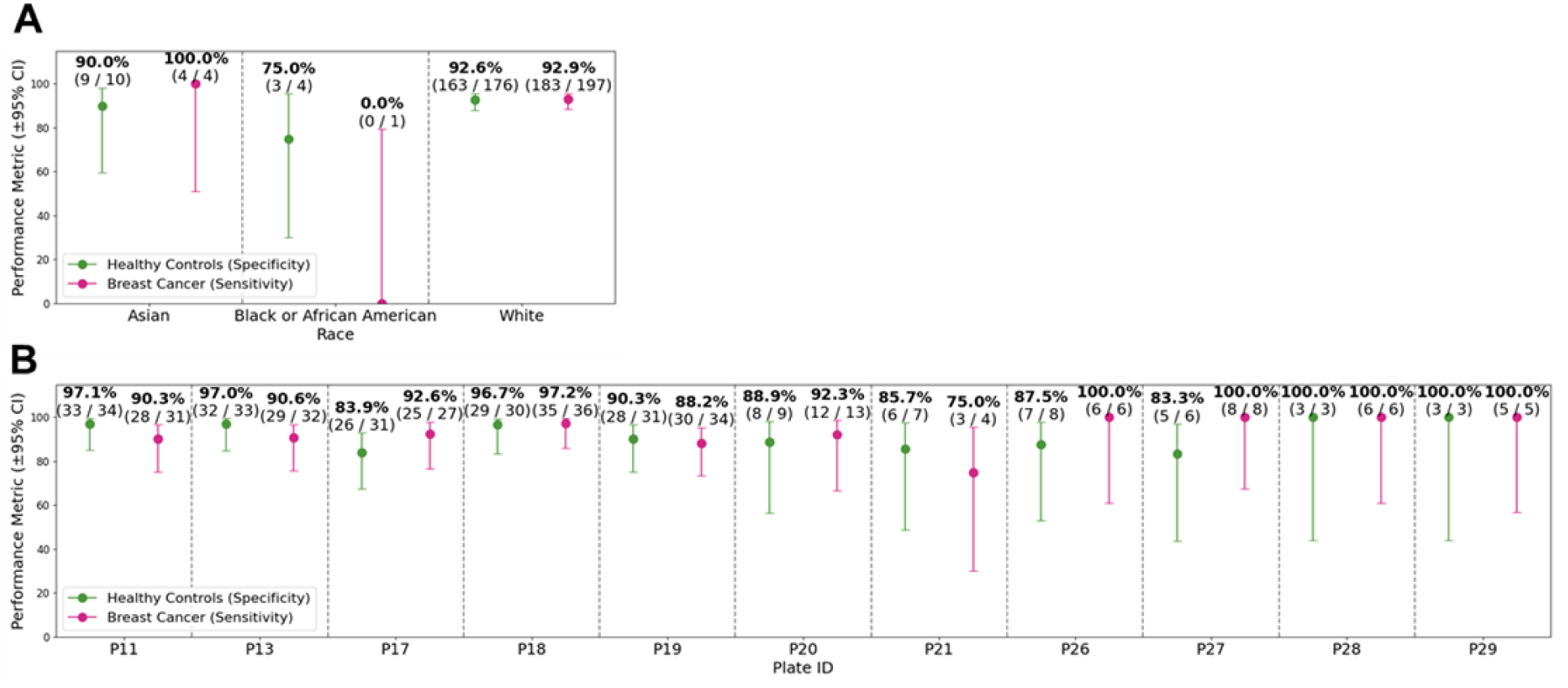
Assessment of confounders based on race and sample preparation batch. To ensure that the training model was not influenced by confounders, we assessed model performance by race (A) and sample preparation batch or by plate (B).

**Supplemental Figure 7.**
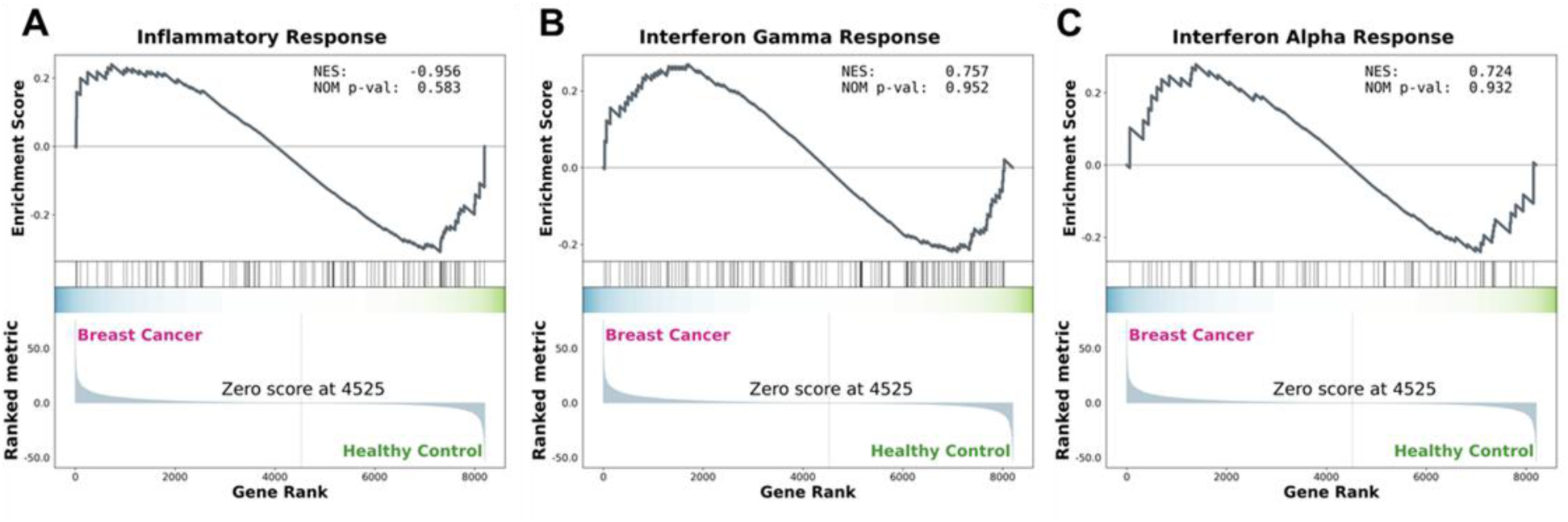
Immune-related pathways are not enriched in breast cancer patients. We evaluated GSEA pathways including inflammatory response (A), interferon gamma response (B), and interferon alpha response (C). Each pathway showed no enrichment in either healthy or breast cancer patient samples.

## References

1. Boyd, N.F., et al. Mammographic density and the risk and detection of breast cancer. N Engl J Med 356, 227–236 (2007).

2. McCormack, V.A. & dos Santos Silva, I. Breast density and parenchymal patterns as markers of breast cancer risk: a meta-analysis. Cancer Epidemiol Biomarkers Prev 15, 1159–1169 (2006).

3. Duffy, S.W., et al. Mammographic density and breast cancer risk in breast screening assessment cases and women with a family history of breast cancer. European journal of cancer 88, 48–56 (2018).

4. Payne, N.R., et al. Breast density effect on the sensitivity of digital screening mammography in a UK cohort. Eur Radiol 35, 177–187 (2025).

5. Mammography Quality Standards Act. (ed. Food and Drug Administration, H.) (2023).

6. Gilbert, F.J., et al. Comparison of supplemental breast cancer imaging techniques-interim results from the BRAID randomised controlled trial. Lancet 405, 1935–1944 (2025).

7. Bakker, M.F., et al. Supplemental MRI Screening for Women with Extremely Dense Breast Tissue. N Engl J Med 381, 2091–2102 (2019).

8. Force, U.S.P.S.T., et al. Screening for Breast Cancer: US Preventive Services Task Force Recommendation Statement. JAMA 331, 1918–1930 (2024).

9. Klein, E.A., et al. Clinical validation of a targeted methylation-based multi-cancer early detection test using an independent validation set. Ann Oncol 32, 1167–1177 (2021).

10. Zajec, M., et al. Mass Spectrometry for Identification, Monitoring, and Minimal Residual Disease Detection of M-Proteins. Clin Chem 66, 421–433 (2020).

11. Barnidge, D.R., et al. Using mass spectrometry to monitor monoclonal immunoglobulins in patients with a monoclonal gammopathy. J Proteome Res 13, 1419–1427 (2014).

12. Banerjee, S. Empowering Clinical Diagnostics with Mass Spectrometry. ACS Omega 5, 2041–2048 (2020).

13 . Horrmann, A., et al. A Plasma-based Deep Proteomic Platform for Early-Stage Breast Cancer Detection. medRxiv, 2025.2009.2022.25336353 (2025).

14. Kistner, F., Grossmann, J.L., Sinn, L.R. & Demichev, V. QuantUMS: uncertainty minimisation enables confident quantification in proteomics. bioRxiv, 2023.2006.2020.545604 (2023).

15. Cox, J., et al. Accurate proteome-wide label-free quantification by delayed normalization and maximal peptide ratio extraction, termed MaxLFQ. Mol Cell Proteomics 13, 2513–2526 (2014).

16. Agarwal, A.B., A; Dudik, M; Langford; Wallach, H. A Reductions Approach to Fair Classification. Proceedings of Machine Learning Research 80(2018).

17. von Euler-Chelpin, M., Lillholm, M., Vejborg, I., Nielsen, M. & Lynge, E. Sensitivity of screening mammography by density and texture: a cohort study from a population-based screening program in Denmark. Breast Cancer Res 21, 111 (2019).

18. Potsch, N., Vatteroni, G., Clauser, P., Helbich, T.H. & Baltzer, P.A.T. Contrast-enhanced Mammography versus Contrast-enhanced Breast MRI: A Systematic Review and Meta-Analysis. Radiology 305, 94–103 (2022).

19. Kelly, K.M., Dean, J., Comulada, W.S. & Lee, S.J. Breast cancer detection using automated whole breast ultrasound and mammography in radiographically dense breasts. Eur Radiol 20, 734–742 (2010).

20. Pereslucha, A.M., Wenger, D.M., Morris, M.F. & Aydi, Z.B. Invasive Lobular Carcinoma: A Review of Imaging Modalities with Special Focus on Pathology Concordance. Healthcare (Basel) 11(2023).

21. Panet, F., et al. Use of ctDNA in early breast cancer: analytical validity and clinical potential. NPJ Breast Cancer 10, 50 (2024).

